# TOP-Plus is a Versatile Biosensor Platform for Monitoring SARS-CoV-2 Antibody Durability

**DOI:** 10.1101/2021.02.03.21251089

**Authors:** Sabrina E. Racine-Brzostek, Mohsen Karbaschi, Christian Gaebler, P.J. Klasse, Jim Yee, Marina Caskey, He S. Yang, Ying Hao, Amy Chadburn, Yuanyuan Shi, Robert Zuk, Michel C. Nussenzweig, Melissa M. Cushing, Zhen Zhao

## Abstract

**Background:** There is a concern that low initial SARS-CoV-2 antibody titers in individuals may drop to undetectable levels within months after infection. Although this may raise concerns over long term immunity, both the antibody levels and avidity of the antibody-antigen interaction should be examined to understand the quality of the antibody response.

**Methods:** A testing-on-a-probe “plus” panel (TOP-Plus) was developed, which included a newly developed avidity assay built into the previously described SARS-CoV-2 TOP assays that measured total antibody (TAb), surrogate neutralizing antibody (SNAb), IgM and IgG on a versatile biosensor platform. TAb and SNAb levels were compared with avidity in previously infected individuals at 1.3 and 6.2 months post-infection in paired samples from 80 COVID-19 patients.

**Results:** The newly designed avidity assay in this TOP panel correlated well with a reference Bio-Layer Interferometry avidity assay (R=0.88). The imprecision of the TOP avidity assay was less than 9%. Although TAb and neutralization activity (by SNAb) decreased between 1.3 and 6.2 months post infection, the antibody avidity increased significantly (P < 0.0001).

**Conclusion:** This highly precise and versatile TOP-Plus panel with the ability to measure SARS-CoV-2 TAb, SNAb, IgG and IgM antibody levels and avidity of individual sera on one sensor can become a valuable asset in monitoring not only SARS-CoV-2-infected patients, but also the status of individuals’ COVID-19 vaccination response.

## Introduction

Although coronavirus disease 2019 (COVID-19), caused by the novel severe acute respiratory syndrome coronavirus 2 (SARS-CoV-2), results in either an asymptomatic or mild infection, it has nonetheless led to crippling levels of morbidity and mortality around the world.*(1)* After the unprecedentedly rapid development of multiple diagnostic nucleic acid tests, serologic testing became the focus in COVID-19 testing. Initially these serologic assays were aimed at identifying individuals with prior exposure to SARS-CoV-2 and potential convalescent plasma donors, as well as supporting epidemiologic studies during the public health emergency.*(2, 3)*

Seroprevalence studies have begun to show a larger degree of SARS-CoV-2 infections than initially reported because of the high prevalence of infected individuals with mild or no symptoms.*(4, 5)* However, lower SARS-CoV-2 IgG antibody levels have been reported in those with mild or no symptoms when compared to those with severe COVID-19.*(6-9)* Furthermore, emerging evidence suggests that SARS-CoV-2 antibodies in some asymptotic carriers may diminish over time to levels below detection.*(10-12)* This decrease in antibody levels over time may include neutralizing SARS-CoV-2 antibodies, which play a vital role in viral clearance.*(9)* Together, this puts into question whether acquired immunity may be short lived and *herd immunity* protection may be less durable than anticipated.*(13)*

While many studies focus on overall antibody titers, other factors are likely equally important in evaluating the humoral antibody response. Binding titers are the products of antibody concentration and average affinity. Avidity can be defined as the strengthening of antibody binding through bi- or multivalency or as the functional affinity of the entire IgG, IgA, or IgM molecule, a net product of the intrinsic paratope-epitope affinity and the valency.*(14)* In the following we use the term *avidity* in the latter sense. Low avidity antibodies are typically produced early in the humoral immune response.*(8, 15)* Over time, with affinity maturation, the intrinsic affinity of the antibody-antigen interaction strengthens, and so does the functional affinity or avidity of bivalent IgG or classes of higher valency.

To evaluate whether these reported weak early antibody responses should be of clinical concern, various assays have emerged to help assess antibody avidity in the evaluation of the SARS-CoV-2 immune response. *(16)* Antibody avidity may be measured in a variety of ways. However, equilibrium binding assays or endpoint titrations may not completely describe all relevant variables. Enzyme-linked immunosorbent assays (ELISA), high performance liquid chromatography (HPLC), capillary electrophoresis or single radial immunodiffusion (SRID) are labor intensive, low-throughput and display low accuracy and precision. Biosensor technologies such as Surface Plasmon Resonance (SPR) and Bio-Layer Interferometry (BLI) have become popular in monitoring the molecular binding between antigen and antibody in a real-time and cost effective manner.*(17)* This study describes a similar, yet novel, approach to evaluating the level and avidity of SARS-CoV-2 receptor-binding domain (RBD) antibodies using a testing-on-a-probe (TOP) plus panel (TOP Plus) which includes a newly developed avidity assay and the previously described SARS-CoV-2 TOP assays (total antibody [TAb], surrogated neutralizing antibody [SNAb]) on a single versatile biosensor platform. This fully automated assay panel was used in this study to evaluate and describe the antibody response and antibody avidity approximately 1 month and 6 months post symptom onset in 80 individuals previously diagnosed with COVID-19. *(18)*

## Materials and methods

### Study participants and Source of Specimen

The study was approved by Rockefeller University (IRB# DRO-1006) and determined to meet exemption requirements by Weill Cornell Medicine Institutional Review Board.

The details of the patient characteristics have been described previously. *(15, 18)*. Eighty out of 87 previously included participants were included in this study. Seven prior participants did not consent to sample sharing. In summary, eligible participants were adults aged 18-76 years and were either diagnosed with SARS-CoV-2 infection by RT-PCR, or were in close contact (e.g., household, coworkers, members of same religious community) with someone who had been diagnosed with SARS-CoV-2 infection by RT-PCR. These study participants had been recruited at the Rockefeller University Hospital in New York from 1 April to 8 May 2020 during the initial screen (approximately 1.3 months after initial infection/onset of symptoms) and returned August 31 through October 16, 2020 for follow up (approximately 6.2 months after the onset of symptoms). Blood samples were collected at the Rockefeller University Hospital, and Weill Cornell Medicine performed the antibody analyses as described below.

### SARS-CoV-2 total antibody and surrogate neutralizing antibody assays

The SARS-CoV-2 total antibody TAb and SNAb assays were used to measure plasma TAb and SNAb antibodies against SARS-CoV-2. Plasma samples were assayed on the fully automated Pylon 3D analyzer (ET HealthCare, Palo Alto, CA), as previously described.*(19, 20)* Briefly, the TAb analysis was performed using a unitized test strip containing wells with predispensed reagents. The TAb reagent contains biotinylated recombinant versions of the SARS-CoV-2 S-Protein RBD as antigens that bind the antibody. The TAb assay measures the overall binding (of all five antibody classes) between SARS-CoV-2 antibodies and the SARS-CoV-2 S-Protein RBD. The assay is based on RBD pre-coated probes and preloaded reagent strips. The SNAb assay is based on the anti-SARS-CoV-2-S-protein antibody-mediated inhibition of the interaction between the ACE2 receptor and the RBD of the S protein. The percentage of ACE2-RBD binding in the SNAb assay is an inverse surrogate for virus-neutralization (SNAb binding inhibition) and previously had been shown to correlate well with both plaque reduction neutralization test (PRNT) and pseudo virus neutralization test (PsV). *(20)*

### SARS-CoV-2 antibody avidity assay

The principle of the SARS-CoV-2 antibody avidity assay is similar to a previously described technology *(20)* that measured SARS-COV-2 antibodies at the tip of an RBC-coated quartz probe and used a biotinylated RBD and a Cy5-Streptavidin conjugate as the signaling elements. However, the calculated relative dissociation rate (dR) allows for avidity testing in this new assay. (Figure 1) In short, a RBD precoated probe is sequentially incubated in microwells containing the sample (to capture SARS-CoV-2 specific antibodies), biotinylated RBD and Streptavidin-Cy5 conjugate along with washes between the incubation steps. After the initial fluorescent signal is measured (Signal_0), the probe with the immobilized immunocomplex enters into repetitive dissociation cycles with multiple incubations in phosphate-buffered saline with Tween^®^ detergent (PBST, pH 7.4) as a dissociation buffer. After each incubation, the fluorescent signal is measured (Signal_t). In the end, a dissociation curve is constructed by plotting the normalized fluorescent signal (Signal_0/Signal_t) over time. The dR (1/s) is calculated by fitting a first order reaction kinetics to the dissociation curve.

**Figure 1.**
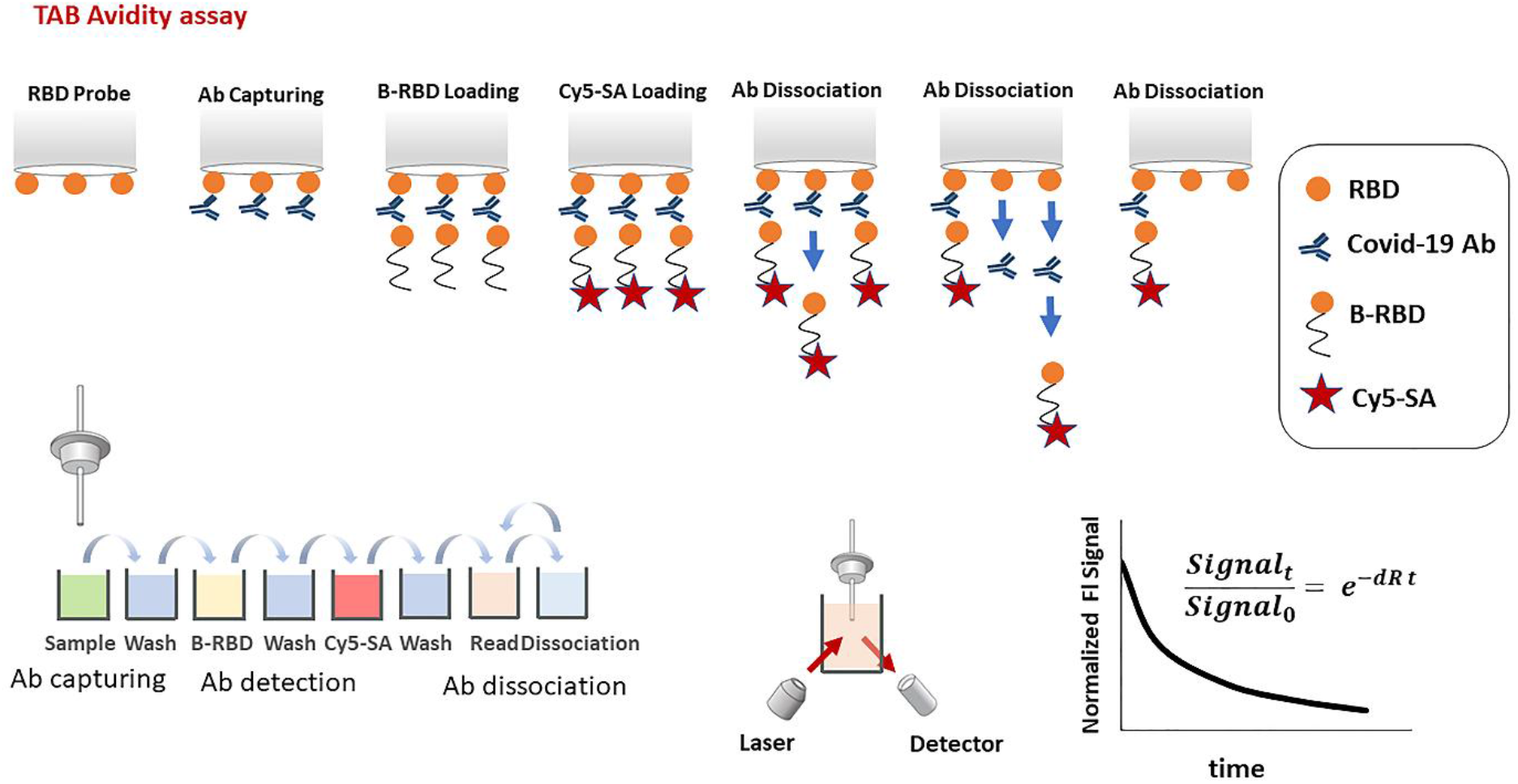
Sars-CoV-2 antibody avidity assay principle. An RBD precoated probe is sequentially incubated in microwells containing 1) sample to capture SARS-CoV-2-specific antibodies 2) wash buffer 3) detection biotinylated RBD 4) wash buffer 5) Streptavidin-Cy5 conjugate. The fluorescent signal is then measured (Signal_0). After that, probe with the immobilized immunocomplex goes into repetitive dissociation cycles by multiple incubations in PBST as dissociation buffer. After each incubation, fluorescent signal is measured (Signal_t). At the end of measurement, dissociation curve is constructed by plotting the normalized fluorescent signal (Signal_0/Signal_t) over time. The relative dissociation rate (dR) is then calculated by fitting a first order reaction kinetics to the dissociation curve.

The dissociation profile represents the rate of antibody dissociation from the RBD coated probe. For an accurate measurement of antibody dissociation rate, a limited but adequate amount of antibody shall be loaded on the probe surface. Loading a high amount of antibody, determined by getting a high initial fluorescent signal (Signal_0) over a certain threshold, causes the formation of a packed multi-layer antibody construct on the probe surface. This leads to an inaccurate measurement as antibodies in a packed adsorbed layer cannot freely dissociate. Therefore, antibody packing density affects the dissociation measurement. On the other hand, sensitive measurements require adequate antibody loading determined by the initial fluorescent signal above a certain level. As such, the proper antibody loading must be within a proper range for accurate measurement. The appropriate initial fluorescent signal (which verifies an optimal antibody loading) was practically determined via a titration study to be in the range of 20-615RFU, as discussed below in the SARS-CoV-2 antibody avidity analytical validation section. Samples with high antibody concentrations must be diluted for measurement. The dilution factor was determined by measuring the initial fluorescent signal to fall within this proper signal range.

Of note, a lower dissociation rate reflects both affinity maturation and multivalent binding development. Either a higher intrinsic binding strength of an antibody paratope to RBD or addition of binding paratopes to the antibody structure results in a higher binding strength and a lower relative dissociation rate of a COVID-19 antibody-RBD pair.

### Precision and interference

The imprecision was determined by running the high and low level of pooled patient samples five times per day for five days. The imprecision of the avidity assay was determined by coefficient of variation (CV).

Specificity or cutoff values were not determined as the avidity is measured only when TAb is positive.*(15, 18)* As explained in the validation results that follow, the ideal fluorescent signal range as an indication for optimal antibody loading is 20-615 RFU. As a negative TAb has a Signal_0 below 20 RFU in the clinical validation, it does not fall within this ideal range of measurement.

Common endogenous interferences were tested on the avidity of two SARS-CoV-2 model purified antibodies in pooled SARS-CoV-2 negative serum (SinoBiological 40150-D001; Absolute Antibody Ab01680-10). Serum was spiked with 0.1mg/ml Biotin, 0.2mg/ml Bilirubin, 5mg/ml Hemoglobin or 2mg/ml Triglyceride and avidity was measured. Recovery was reported as the percentage ratio of dR measurements in spiked over unspiked serum. Measurements were performed in quadruplicate for assay precision.

### Bio-Layer Interferometry

Gator (Gator Bio, Palo Alto, CA, USA) was used to measure avidity of 12 different purified COVID-19 antibodies. Gator koff (dissociation constant) measurement was performed in a buffer of 10 ug/ml antibody with 0.2% BSA and 0.02% Tween 20.

## Results

### SARS-CoV-2 antibody avidity analytical validation

The SARS-CoV-2 antibody avidity assay was initially tested with 12 different purified antibodies against SARS-CoV-2 purchased from different vendors. Human sera from SARS-CoV-2 negative patients were spiked with these antibodies and measured for avidity. At first, a titration study was performed to determine the proper range of antibody loading. 30 ug/ml antibody samples were diluted in series and applied for dR measurement. Samples with initial fluorescent signal (Signal_0) in the range of 20-615 relative fluorescence unit (RFU) showed consistent dR values, independent of the signal or concentration level. We considered this fluorescent signal range as an indication for optimal antibody loading. Samples with a high fluorescent signal (Signal_0 > 615RFU) were diluted accordingly for measurement. Samples with a low fluorescent signal (Signal_0 <20 RFU) were identified as unmeasurable.

Figure 2A demonstrates the dissociation profiles of five different antibodies over time. Antibodies of varying RBD binding strength displayed different relative dissociation rates and therefore, different dissociation profiles. The relative dissociation rates were measured at proper antibody loading concentrations (0.06-30ug/ml, varying for different antibody) and the avidity measurement was found to be concentration (and therefore fluorescent signal) independent, as long as the initial fluorescent signal (Signal_0) is in the proper range (Fig 2B).

**Figure 2.**
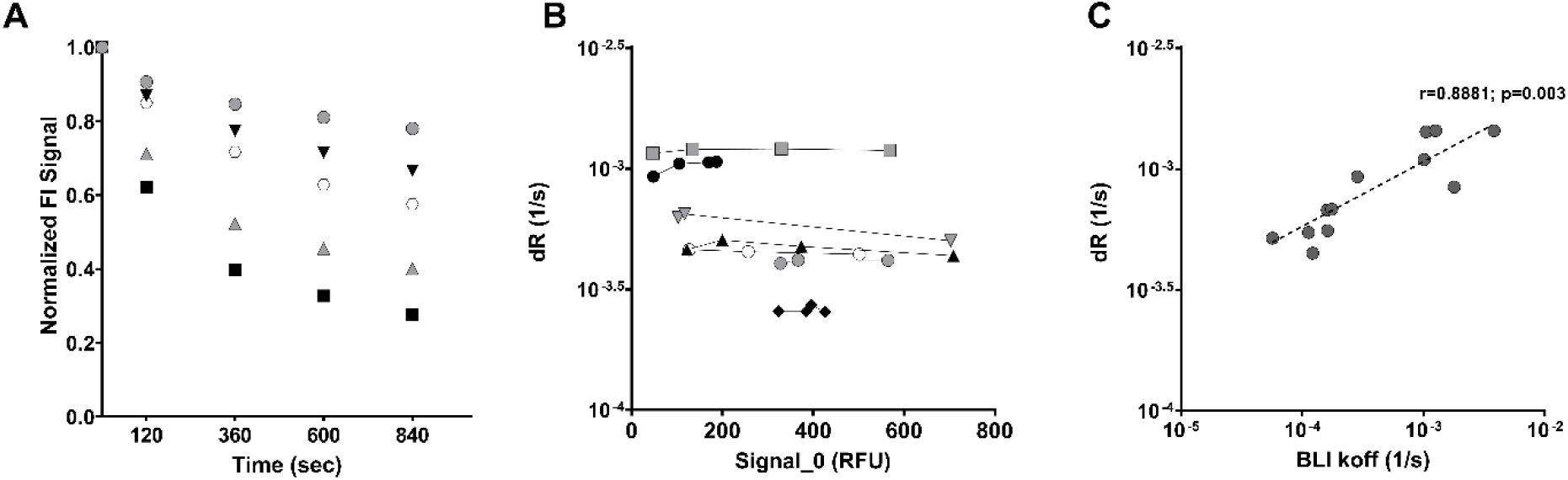
Sars-CoV-2 antibody avidity assay characterization with model COVID-19 purified antibodies. A) Dissociation curve measurement of COVID-19 negative human serum spiked with five different COVID-19 antibodies. B) Relative dissociation rate measurement at varying levels of Signal_0. COVID-19 negative human serum were spiked with COVID-19 antibodies at different levels and measured. C) Correlation between the Sars-CoV-2 avidity assay (dissociation rate measurement) and the Bio-Layer Interferometry (BLI) measurement (koff). Correlation between the two assays were assessed by Spearman correlation coefficient.

BLI is a well-established technique in avidity measurement *(21, 22)*. To further validate the performance of TOP-Plus’ avidity assay, Gator was used as a BLI reference method to measure avidity of 12 different purified COVID-19 antibodies. Avidity of these antibodies were also measured with the TOP-Plus avidity assay using COVID-19 negative pooled serum spiked with one of these antibodies at a proper loading level. It was found that the TOP-Plus avidity assay measurements correlated well (r=0.88) with the Gator measurements (Figure 2C).

The TOP-Plus avidity assay was tested with common endogenous immunoassay interferences. Avidity of two SARS-CoV-2 model purified antibodies first were measured in pooled SARS-CoV-2 negative serum. This was followed with spiking either 0.1mg/ml Biotin, 0.2mg/ml Bilirubin, 5mg/ml Hemoglobin or 2mg/ml Triglyceride and the measurement of antibody avidity in presence of each potential interferents. The TOP-Plus avidity assay displayed no interference from the listed components up to the tested concentrations. (Table 1)

**Table 1.** Interference and precision study of the avidity assay. Relative dissociation rate of two different SARVS-CoV-2 antibodies is measured in COVID-19 negative serum spiked with different levels of Biotin, Triglyceride, Bilirubin and Hemoglobin. Precision was calculated from quadruplicate measurements.

| Matrix | Human Serum | Human Serum with: |  |  |  |
| --- | --- | --- | --- | --- | --- |
|  |  | 0.1 mg/mL Biotin | 2mg/mL Triglyceride | 0.2mg/mL Bilirubin | 5mg/mL Hemoglobin |
| dR (1/s) | 4.94 E-04 | 5.08 E-04 | 4.53 E-04 | 5.35 E-04 | 5.15 E-04 |
| CV% | 4.1 | 3.7 | 8.5 | 10.8 | 5.8 |
| Recovery % | 100 | 103 | 92 | 108 | 104 |

B. SARS-CoV-2 Spike antibody, Human MAB IgG koff (BLI): 1.01E-03 1/s
| Matrix | Human Serum | Human Serum with: |  |  |  |
| --- | --- | --- | --- | --- | --- |
|  |  | 0.1 mg/mL Biotin | 2mg/mL Triglyceride | 0.2mg/mL Bilirubin | 5mg/mL Hemoglobin |
| dR (1/s) | 8.28 E-04 | 8.38 E-04 | 8.00 E-04 | 8.12 E-04 | 8.91 E-04 |
| CV% | 2.1 | 2.1 | 6.6 | 3.8 | 4.3 |
| Recovery % | 100 | 101 | 97 | 98 | 108 |

The imprecision was determined by running the high and low level of pooled patient samples (n=5-10) five times per day on five different days. The imprecision of the avidity assay CV was found to be < 9%. The stability of samples at 2-4°C refrigerated conditions was was at least 5 days (variation: < 8%).

### Determination of concentration independent range in clinical specimen

As mentioned above, in order to measure an accurate antibody relative dissociation rate, only sufficient amounts of antibody should be loaded onto the probe surface as high amounts of antibody cause the formation of a packed multi-layer antibody construct on the probe surface. These antibodies in the packed adsorbed layer cannot freely dissociate and the antibody packing density affects the dissociation measurement. To evaluate and minimize this potential source of artifact associated with these label-free methods, dilution studies were performed using four specimens with high TAb measurements (in the range 1171-7494 RFU and plotted against the relative disassociation rate. (Figure 3A). Based on these studies, it was determined that the concentration-independent range for this assay is between 20 and 615 TAb RFU. (Figure 3B) Therefore, any specimen with a TAb greater than 615 RFU are first diluted into the 20-615 RFU range prior to determining the relative dissociation rate.

**Figure 3.**
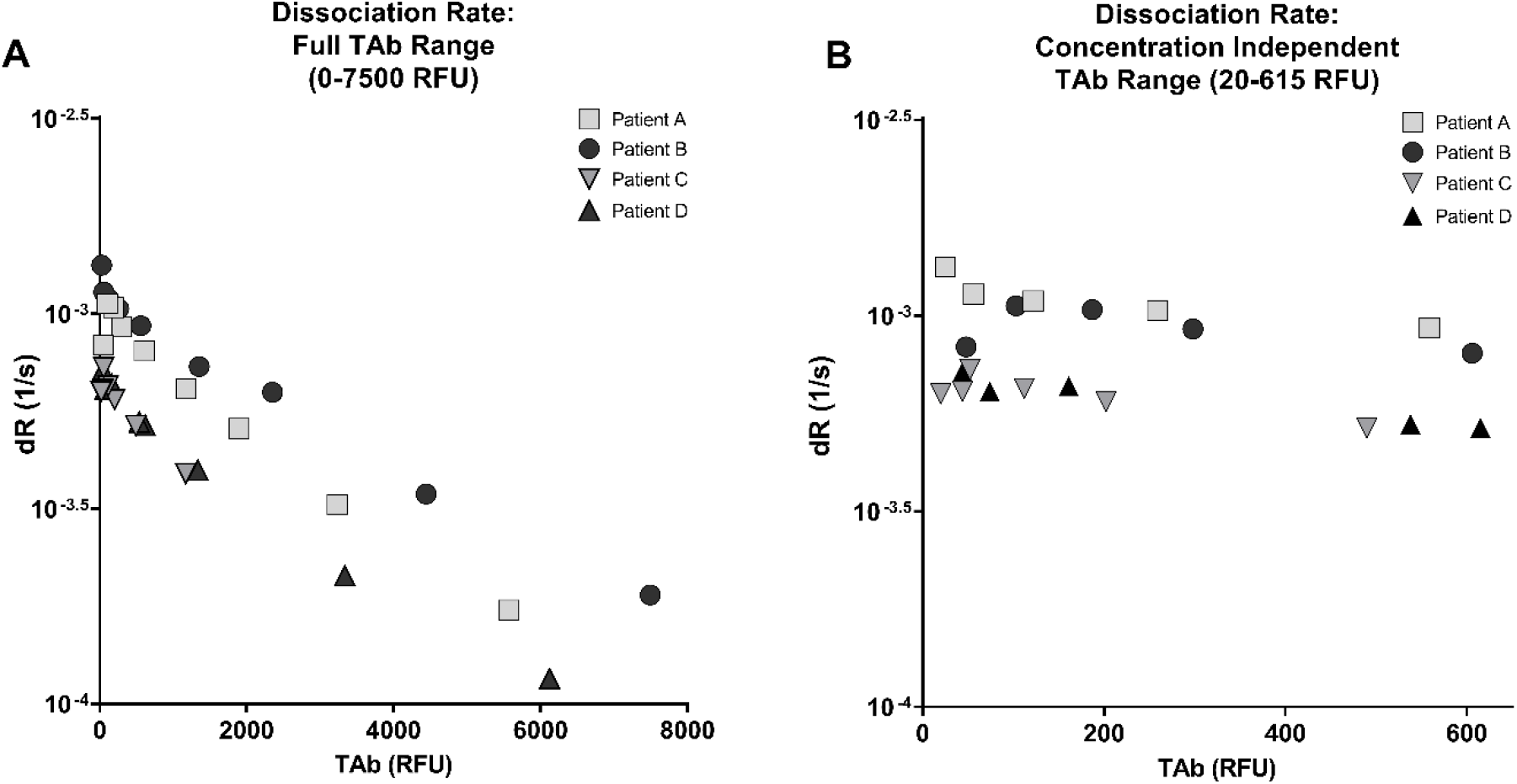
Determination of concentration independent range. A) Dilution studies were performed using four specimen with high TAb measurements, (1171-7494 undiluted RFU) and plotted against the disassociation rate. B) The dissociation rate was independent of TAb concentration between 20 and 615 TAb RFU.

### Evaluation of clinical utility: SARS-CoV-2 antibody dynamics during early convalescence

Serum from 80 COVID-19 individuals *(15, 18)* were evaluated approximately 1.3 months and again 6.2 months after confirmed or suspected time of SARS-CoV-2 infection. TAb levels in the majority of individuals decreased over time with a difference in means of -0.4747 RFU (p=0.0042; Figure 4A-C). This was reflective of the decrease in IgG and IgM levels measured using the same TOP biosensor during this time period which were previously described in this same cohort in prior studies. *(15, 18, 19)* Also, neutralization activity decreased during this time period, as reflected by the lower SNAb binding activity, with a difference in means of 18.81 (%B/B0; p< 0.0001; Figure 4D-F)). In contrast, the avidity increased, as indicated by a significant decrease in mean dR: 9.685×10^−4^/s at 1.3 months post infection to 5.83×10^−4^/s at 6.2 months post infection (p < 0.0001; Figure 4G-I).

**Figure 4.**
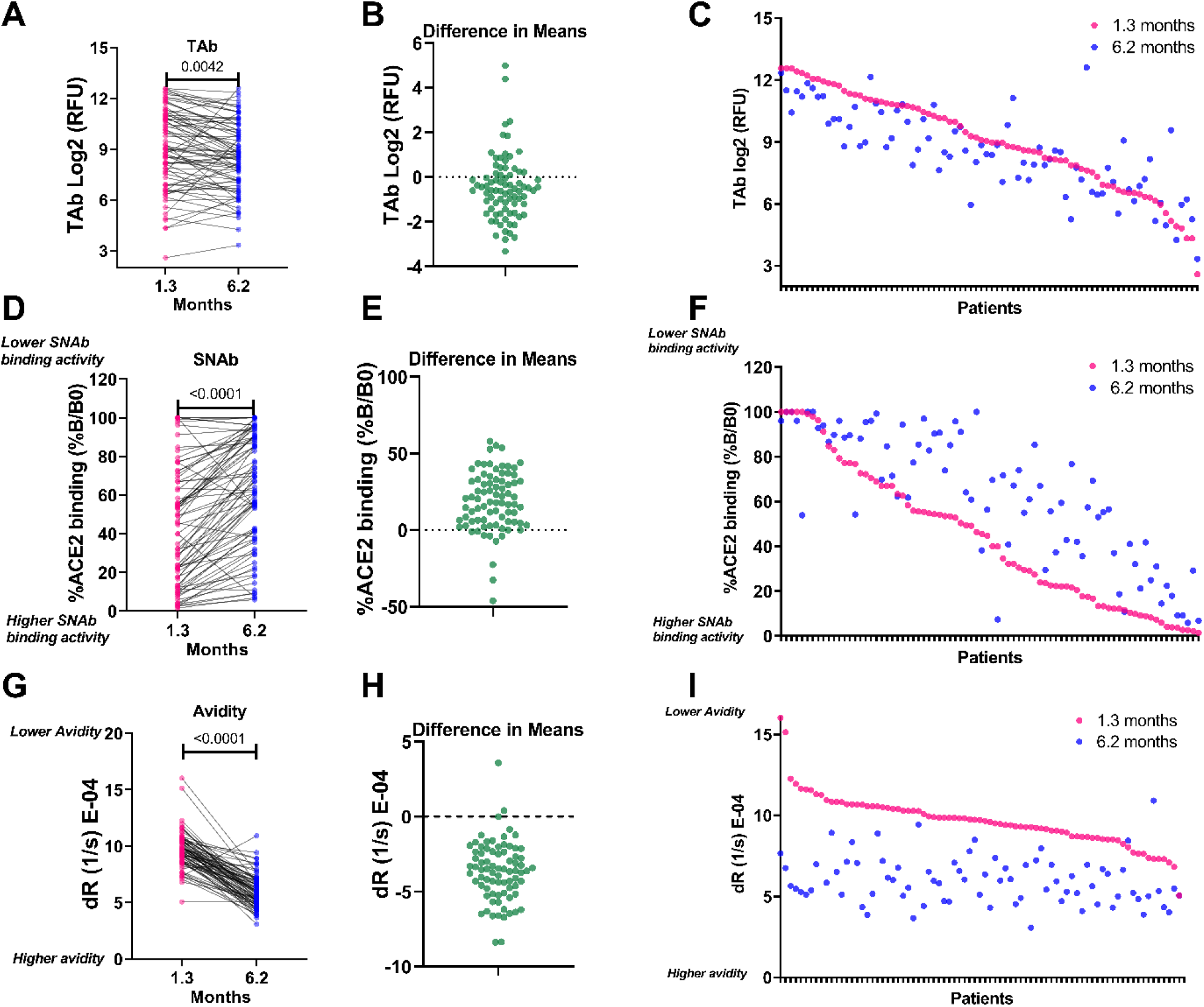
SARS-CoV-2 antibody dynamics during early convalescence (1-6 months post infection) Eighty COVID-19 positive individuals were evaluated approximately 1.3 months and again 6.2 months after confirmed or suspected time of SARS-CoV-2 infection. TAb (A-C) and SNAb (D-F) levels in the majority of individuals decreased over time. In contrast, the relative dissociation rate (reflecting avidity) increased during this time period (G-I). Panels B, E, H respectively demonstrate the change in TAb, SNAb and relative dissociation rate between 1.3 and 6.2 months post infection. Please note that percentage of ACE2-RBD binding in the SNAb assay is an *inverse* surrogate for virus-neutralization (SNAb binding inhibition) and previously had been shown to correlate well with PRNT and PsV. The t-test was performed between numerical variables. A p-value < 0.05 was considered statistically significant.

## Discussion

Antibody avidity testing is not a new concept in the evaluation of an antibody response to infection or vaccination. It has also played a role in other important applications, as in the generation of efficacious therapeutic antibodies. *(23)* Typically, antibodies generated early in a primary infection bind weakly to their respective antigen and exhibit low avidity or functional affinity. However, overall avidity towards an antigen(s) increases as the response matures through somatic hypermutation particularly of the variable loops of antigen-binding sites of B cell receptors and selective survival in the germinal center. *(24, 25)* As it typically increases over time and is an indicator of a more mature antibody response, antibody avidity could be applied in assessing the efficacy of COVID-19 vaccination, immunity to SARS-CoV-2 and screening donors for convalescent plasma antibody therapies.

Studies have tried to explain the variability of SARS CoV-2 antibody response by focusing on antibody avidity.*(26)* In this current study we monitored avidity by measuring the relative rate of SARS-CoV-2 specific antibodies dissociation from RBD in addition to TAb and SNAb. This assay allows the assessment of the antibodies’ strength in binding to the virus. The relative dissociation rate inversely associates with the average antibody’s residence time at the epitope. Antibodies with lower relative dissociation rate values tightly bind to RBD and therefore may be more efficient in clearing the virus and neutralizing infectivity, i.e., blocking the entry into target cells.*(27)*

Antibody affinity reflects the rate constants of association and dissociation of an antibody with its target antigen (K_D_ (M)= k_off_ (1/s) /k_on_ (1/Ms). In many serological applications, measurement of antibody-antigen interactions becomes a complicated process. Therefore, the most common approach is to disrupt the antibody-antigen binding by chaotropic agents (such as urea). The avidity is then assessed by measuring the change in the degree of release of antibody from the antigen by the chaotropic agent.*(28, 29)* As a result, the assessed avidity of antibody depends on its resistance to the chaotropic agent and may not truly represent the avidity of antibody toward the antigen.*(14)*

The TOP-Plus avidity assay presented here measures the relative rate of dissociation of SARS-CoV-2 antibodies from the RBD antigen in plasma. However, this assay distinguishes itself from others in that it does not apply a chaotropic reagent. Therefore, the measured dR values better reflect the natural relative dissociation rate of antibodies from their target antigen than the conventional approaches where chaotropes may alter the native structure of the antigen or antibody. *(14)*

Previously no one assay could evaluate total antibody levels, individual IgM and individual IgG levels as well as avidity. The new TOP-Plus biosensor panel comprises five assays, allowing for TAb, SNAb, IgG and IgM levels as well as avidity testing on the same platform using the same biosensor principles with specific application applied for each assay. For example, as described earlier, the probe was able to assess the overall decreasing trend in TAb and SNAb .(Figure 4) In addition, as described previously, IgG and IgM also display decreasing trends in SARS-CoV-2 antibody levels.*(15)* However, despite these decreasing antibody levels, the overall avidity of the SARS-CoV-2 antibodies increase (Figure 4C). This allows for clinical assurance of an adequate humoral immune response to the SARS-CoV-2 as the patient convalesces without the need of multiple blood draws and testing. With the ongoing world-wide roll out of COVID-19 vaccinations, such a panel could also play a major role in monitoring the vaccination response.

A limitation of the study is that only convalescent serum specimens were used for evaluation. Current data cannot speak to the acute phase of infection as avidity maturation occurs. This will require further studies in order to determine the utility of this new avidity assay in acutely ill patients. As it is a newly evolved virus, it would be expected that the antibody avidity for SARS-CoV-2 antigens during primary infection would be weak and this avidity would increase over time. However, during the acute stages of infection, IgM could precede the IgG response. It could be postulated that the overall avidity may display an initial spike during the acute stage of infection due to the multimeric structure of the IgM antibody and may mask the primary infection’s expected weaker avidity.

In conclusion, this TOP-Plus biosensor panel is a versatile sensing platform with high precision and an ability to measure SARS-CoV-2 TAb, SNAb, individual IgG and IgM antibody levels along with the antibody’s long-term avidity. This combination of all-in-one testing will be a valuable asset in monitoring not only convalescent COVID-19 patients, but also the status of individuals’ COVID-19 vaccination response.

## Data Availability

An anonymized dataset and data analysis code may be available upon application to the corresponding author and institutional review as per Weill Cornell Medicine data sharing policies

## Abbreviation list

COVID-19: Corona Virus Disease-2019
SARS-CoV-2: Severe Acute Respiratory Syndrome Coronavirus 2
RT-PCR: real-time polymerase chain reaction
RBD: receptor-binding domain
TOP: testing-on-a-probe
HPLC: high performance liquid chromatography
SRID: single radial immunodiffusion
SPR: Surface Plasmon Resonance
BLI: Bio-Layer Interferometry
RFU: relative fluorescence unit
dR: relative dissociation rate
TAb: total antibody
SNAb: surrogate neutralizing antibody
ELISA: Enzyme-linked immunosorbent assays
PRNT: plaque reduction neutralization test
PsV: pseudo virus neutralization test

## Acknowledgments

We thank all study participants who devoted time to our research; Drs. Barry Coller and Sarah Schlesinger, the Rockefeller University Hospital Clinical Research Support Office and nursing staff. This work was supported by a COVID-19 research grant from Weill Cornell Medicine (M.C.), Weill Cornell Medicine Translational Research Program of the Department of Pathology and Laboratory Medicine at Weill Cornell Medicine (Z.Z.). NIH grants P01 AI 110657 and R01 AI36082 (P.J.K.) and NIH grant P01-AI138398-S1 and 2U19AI111825 (M.C.N.). C.G. was supported by the Robert S. Wennett Post-Doctoral Fellowship, in part by the National Center for Advancing Translational Sciences (National Institutes of Health Clinical and Translational Science Award program, grant UL1 TR001866), and by the Shapiro-Silverberg Fund for the Advancement of Translational Research. M.C.N. is a Howard Hughes Medical Institute Investigators.

## Author Contributions

All authors confirmed they have contributed to the intellectual content of this paper and have met the following 4 requirements: (a) significant contributions to the conception and design, acquisition of data, or analysis and interpretation of data; (b) drafting or revising the article for intellectual content; (c) final approval of the published article; and (d) agreement to be accountable for all aspects of the article thus ensuring that questions related to the accuracy or integrity of any part of the article are appropriately investigated and resolved.

JY performed the experiments. YH helped collect data. SRB wrote the manuscript, generated figures and aided in the review of the investigational findings. ZZ oversaw the project, including conceptualization, interpretation of the data, statistical analysis and writing of the manuscript. SY helped edit the manuscript and interpretation of the data. AC and MMC helped review the manuscript. CG collected specimens, analyze the patient data and review of the manuscript. RZ oversaw the methodology for the TOP-Plus method development and conceptualization of the project. MK performed formal analysis for the TOP assay development, provided analytical data analysis and helped edit the manuscript.

PJK helped with editing the manuscript and performed a formal review. MC collected specimens, analyze the patient data and review of the manuscript. MCN collected specimens, analyze the patient data and review of the manuscript. YYS helped with conceptualization of the project and design the experiments.

## References

1. Gao F, Zheng KI, Wang XB, Sun QF, Pan KH, Wang TY, et al. Obesity is a risk factor for greater covid-19 severity. Diabetes Care 2020;43:e72–e4.

2. Lisboa Bastos M, Tavaziva G, Abidi SK, Campbell JR, Haraoui LP, Johnston JC, et al. Diagnostic accuracy of serological tests for covid-19: Systematic review and meta-analysis. BMJ 2020;370:m2516.

3. Moura DTH, McCarty TR, Ribeiro IB, Funari MP, Oliveira P, Miranda Neto AA, et al. Diagnostic characteristics of serological-based covid-19 testing: A systematic review and meta-analysis. Clinics (Sao Paulo) 2020;75:e2212.

4. Sood N, Simon P, Ebner P, Eichner D, Reynolds J, Bendavid E, Bhattacharya J. Seroprevalence of sars-cov-2-specific antibodies among adults in los angeles county, california, on april 10-11, 2020. JAMA 2020;323:2425–7.

5. Havers FP, Reed C, Lim T, Montgomery JM, Klena JD, Hall AJ, et al. Seroprevalence of antibodies to sars-cov-2 in 10 sites in the united states, march 23-may 12, 2020. JAMA Intern Med 2020.

6. Xie J, Ding C, Li J, Wang Y, Guo H, Lu Z, et al. Characteristics of patients with coronavirus disease (covid-19) confirmed using an igm-igg antibody test. J Med Virol 2020.

7. Chen Y, Zuiani A, Fischinger S, Mullur J, Atyeo C, Travers M, et al. Quick covid-19 healers sustain anti-sars-cov-2 antibody production. Cell 2020.

8. Li K, Huang B, Wu M, Zhong A, Li L, Cai Y, et al. Dynamic changes in anti-sars-cov-2 antibodies during sars-cov-2 infection and recovery from covid-19. Nat Commun 2020;11:6044.

9. Zheng Y, Yan M, Wang L, Luan L, Liu J, Tian X, Wan N. Analysis of the application value of serum antibody detection for staging of covid-19 infection. J Med Virol 2020.

10. Patel MM, Thornburg NJ, Stubblefield WB, Talbot HK, Coughlin MM, Feldstein LR, Self WH. Change in antibodies to sars-cov-2 over 60 days among health care personnel in nashville, tennessee. JAMA 2020.

11. Long QX, Tang XJ, Shi QL, Li Q, Deng HJ, Yuan J, et al. Clinical and immunological assessment of asymptomatic sars-cov-2 infections. Nat Med 2020;26:1200–4.

12. Wajnberg A, Amanat F, Firpo A, Altman DR, Bailey MJ, Mansour M, et al. Robust neutralizing antibodies to sars-cov-2 infection persist for months. Science 2020;370:1227–30.

13. Brown TS, Walensky RP. Serosurveillance and the covid-19 epidemic in the us: Undetected, uncertain, and out of control. JAMA 2020;324:749–51.

14. Klasse PJ. How to assess the binding strength of antibodies elicited by vaccination against hiv and other viruses. Expert Rev Vaccines 2016;15:295–311.

15. Gaebler C, Wang Z, Lorenzi JCC, Muecksch F, Finkin S, Tokuyama M, et al. Evolution of antibody immunity to sars-cov-2. Nature 2021.

16. Luo YR, Chakraborty I, Yun C, Wu AHB, Lynch KL. Kinetics of sars-cov-2 antibody avidity maturation and association with disease severity. Clin Infect Dis 2020.

17. Petersen RL. Strategies using bio-layer interferometry biosensor technology for vaccine research and development. Biosensors (Basel) 2017;7.

18. Robbiani DF, Gaebler C, Muecksch F, Lorenzi JCC, Wang Z, Cho A, et al. Convergent antibody responses to sars-cov-2 in convalescent individuals. Nature 2020;584:437–42.

19. Yang HS, Racine-Brzostek SE, Lee WT, Hunt D, Yee J, Chen Z, et al. Sars-cov-2 antibody characterization in emergency department, hospitalized and convalescent patients by two semi-quantitative immunoassays. Clin Chim Acta 2020;509:117–25.

20. Yang HS, Racine-Brzostek SE, Karbaschi M, Yee J, Dillard A, Steel PAD, et al. Testing-on-a-probe biosensors reveal association of early sars-cov-2 total antibodies and surrogate neutralizing antibodies with mortality in covid-19 patients. Biosensors and Bioelectronics 2021;178:113008.

21. Sultana A, Lee JE. Measuring protein-protein and protein-nucleic acid interactions by biolayer interferometry. Curr Protoc Protein Sci 2015;79:19 25 1–19 25 6.

22. Kumaraswamy S, Tobias R. Label-free kinetic analysis of an antibody-antigen interaction using biolayer interferometry. Methods Mol Biol 2015;1278:165–82.

23. Sassi AB, Nagarkar R, Hamblin P. Chapter 9 - biobetter biologics. In: Singh M, Salnikova M, editors. Novel approaches and strategies for biologics, vaccines and cancer therapies San Diego: Academic Press; 2015. p. 199–217.

24. Shlomchik MJ. Chapter 22 - selection during antigen-driven b cell immune responses: The basis for high affinity antibody. In: Honjo T, Alt FW, Neuberger MS, editors. Molecular biology of b cells Burlington: Academic Press; 2004. p. 339–48.

25. Merlo LMF, Mandik-Nayak L. Chapter 3 - adaptive immunity: B cells and antibodies. In: Prendergast GC, Jaffee EM, editors. Cancer immunotherapy (second edition) San Diego: Academic Press; 2013. p. 25–40.

26. Bauer G. The variability of the serological response to sars-corona virus-2: Potential resolution of ambiguity through determination of avidity (functional affinity). J Med Virol 2020.

27. Klasse PJ. Neutralization of virus infectivity by antibodies: Old problems in new perspectives. Adv Biol 2014;2014.

28. Benner SE, Patel EU, Laeyendecker O, Pekosz A, Littlefield K, Eby Y, et al. Sars-cov-2 antibody avidity responses in covid-19 patients and convalescent plasma donors. J Infect Dis 2020;222:1974–84.

29. Xiang J, Yan M, Li H, Liu T, Lin C, Huang S, Shen C. Evaluation of enzyme-linked immunoassay and colloidal gold-immunochromatographic assay kit for detection of novel coronavirus (sars-cov-2) causing an outbreak of pneumonia (covid-19). medRxiv 2020:2020.02.27.20028787.

